# On the dependence of the critical success index (CSI) on prevalence

**DOI:** 10.1101/2023.12.03.23299335

**Authors:** Gashirai K. Mbizvo, Andrew J. Larner

**Affiliations:** Pharmacology and Therapeutics, Institute of Systems, Molecular and Integrative Biology, University of Liverpool, United Kingdom; Liverpool Centre for Cardiovascular Science, University of Liverpool and Liverpool Heart & Chest Hospital, Liverpool, United Kingdom; Cognitive Function Clinic, The Walton Centre NHS Foundation Trust, Liverpool, United Kingdom

**Keywords:** Bayes formula, Binary classification, Critical success index, F measure, prevalence

## Abstract

Recently the critical success index (CSI) has been increasingly discussed and advocated as a unitary outcome measure in various clinical situations where large numbers of true negatives may influence the interpretation of other more traditional outcome measures such as sensitivity and specificity, or when unified interpretation of positive predictive value (PPV) and sensitivity (Sens) is needed. The derivation of CSI from measures including PPV has prompted questions as to whether and how CSI values may vary with disease prevalence (P), just as PPV estimates are dependent on P, and hence whether CSI values are generalizable between studies with differing prevalences. As no detailed study of the relation of CSI to prevalence has been undertaken hitherto, the dataset of a previously published test accuracy study of a cognitive screening instrument was reinterrogated to address this question. Three different methods were used to examine the change in CSI across a range of prevalences, using both Bayes formula and equations directly relating CSI to Sens, PPV, P, and to test threshold (Q). These approaches showed that, as expected, CSI does vary with prevalence, but the dependence differs according to the method of calculation adopted. Bayesian rescaling both Sens and PPV generates a concave curve, suggesting that CSI will be maximal at a particular prevalence which may vary according to the particular dataset.

## 1. Introduction

Many measures may be derived from the data cells in a 2x2 contingency table.^1^ Choosing the optimal measure(s) to describe the outcomes of a study may be dependent upon the nature of the available dataset.

For datasets with very large numbers of true negative (TN) outcomes in the base data, as seen for example using routine epilepsy data,^2^ indices such as specificity (Spec), negative predictive value (NPV) and overall classification accuracy (Acc), which all feature TN values in both numerator and denominator, may be very high, indeed approaching values of 1. This is because the numbers of TN may approach the total number of observations (N), and hence swamp the values of the other cells of the 2x2 contingency table, namely true positive (TP), false positive (FP), and false negative (FN).

This circumstance makes it difficult to rank the diagnostic accuracy of the corresponding case-ascertainment algorithms based on Spec, NPV, or Acc, as the figures are all similarly high.^3^ In conditions such as dementia,^4^ motor neurone disease,^5^ and epilepsy,^2^ systematic reviews of the diagnostic accuracy of routine data indicate that the original studies published have largely measured positive predictive value (PPV) and Sens without measuring Spec or NPV. This is because finding true negative cases in the community to verify an absent diagnostic code in a routine dataset is a challenge for researchers, who often only have permission to study populations that have been positively coded with the disease in question. Making a judgment on the optimal case-ascertainment algorithm for a particular condition based on either PPV and Sens is challenging because PPV and Sens tend to have an inverse relationship,^6^ so it is difficult to know which measure to prioritise to best indicate accuracy.

There are other examples in clinical medicine where large numbers of TN may complicate the interpretation of more traditional measures such as PPV and Sens, including National Institute for Clinical Excellence criteria for 2-week-wait suspected brain and CNS cancer referrals,^7^ polygenic hazard scores,^8^ and in the evaluation of cognitive screening instruments.^9^ Accordingly, as we have previously indicated, a metric is needed which eschews TN and combines PPV and Sens. As we are not aware of such a metric currently in common use in medicine, we have proposed use of the critical success index (CSI) for this purpose. This measure, which has been intermittently reinvented over the last century, has been variously known as the ratio of verification in the context of forecasting tornadoes,^10^ and subsequently as the Jaccard index or similarity coefficient (J),^11^ the threat score,^12^ the Tanimoto index,^13^ CSI,^14^ and most recently as F*.^15,16^

In terms of the base data of the 2x2 contingency table:

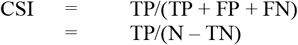

CSI may also be expressed in terms of PPV and Sens:

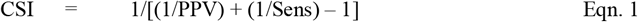

We have demonstrated the advantages of using CSI to complement traditional diagnostic accuracy measures using real-word data in several conditions.^3,9,17^ A question often raised about CSI concerns how its values relate to prevalence, P, the probability of a positive diagnosis. It is well-known that values of PPV vary with P, hence are sensitive to class imbalance and may therefore not be generalizable between studies.^18^ Since, as shown in Eqn.1, CSI may be expressed in terms of PPV, a similar expectation will hold for CSI. Likewise, following from Eqn.1, it may be asked whether CSI values track predominantly with Sens or PPV and whether this changes with P.

Here we initially address two possible methods to illustrate the dependence of CSI on P, as previously suggested:^17^ firstly using Bayes formula to recalculate PPV and then to recalculate CSI (hence a two-step method); and secondly using equations in which CSI is expressed directly in terms of Sens, PPV, P, and the test threshold or probability of a positive test, denoted Q. In addition, we introduce a third method in which Sens is also rescaled, by using Bayes formula to recalculate NPV and hence Sens. This then allows CSI values to be recalculated using both rescaled PPV and Sens.

## 2. Materials and methods

### 2.1 Dataset

The dataset from a screening test accuracy study^19^ of a cognitive screening instrument, the Mini-Addenbrooke’s Cognitive Examination (MACE),^20^ was re-examined. In this study, MACE was administered to consecutive patient referrals (N = 755) to a dedicated cognitive disorders clinic located in a secondary care neurosciences centre. Subjects gave informed consent and the study was approved by the institute’s committee on human research (Walton Centre for Neurology and Neurosurgery Approval: N 310).

In this cohort, 114 patients received a final criterial diagnosis (DSM-IV) of dementia (P = 0.151).^19^ The original analysis of the dataset established the optimal MACE cut-off for the diagnosis of dementia to be ≤20/30 (calculated from the maximal value for the Youden index), where TP = 104, FP = 188, FN = 10, and TN = 453. Hence, at this cut-off, Sens = 0.912, Spec = 0.707, PPV = 0.356, and NPV = 0.978.

From these base data, values of CSI across a range of P values (0.1 to 0.9, in 0.1 increments) were calculated using three different methods.

### 2.2 Method 1: CSI recalculated via Bayes formula for PPV

As Sens and Spec are relatively impervious to change in P, being strictly columnar ratios in the 2x2 contingency table, PPV may be recalculated for different values of P using Bayes formula:

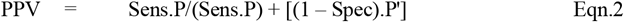

where P’ = (1 – P). Using the base data (Sens = 0.912, Spec = 0.707) values of PPV were calculated for P values ranging from 0.1 to 0.9.

The second step in this method used the recalculated PPV values at different prevalences to recalculate CSI values according to its relation to PPV and Sens (Eqn. 1).

Hence this approach requires the sequential application of Eqn.2 and Eqn.1 to the base data. Results were displayed in a table and graphically.

### 2.3 Method 2: CSI recalculated via its relation to Sens, PPV, P, and Q

The dependence of CSI on P, the probability of a positive diagnosis, may be directly expressed in terms of Sens, PPV, P, and test threshold, the probability of a positive test (Q):^1^

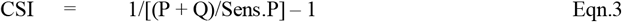

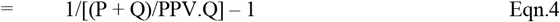

Hence, the dependence of CSI on P may be addressed by calculating its value for different values of P at chosen values of Q. Q ranges from 0-1, where Q = 0 equates to a test threshold at which there are no positives (neither TP nor FP), and Q = 1 equates to a threshold at which there are no negatives (neither TN nor FN). When Q = 0.5, in a balanced data set (P = 0.5) there are equal numbers of false positives and false negatives.

Using the base data (Sens = 0.912, PPV = 0.356), values of CSI were calculated for P values ranging from 0.1 to 0.9 to illustrate the dependence of CSI on P. Three conditions were examined: Q = 0.1 (very few false positives); Q = 0.5 (equal numbers of false positives and false negatives, if the dataset was balanced); and Q = 0.9 (very few false negatives).

Hence this approach requires the application of either Eqn.3 or Eqn.4 to the base data. Results were displayed in tables and graphically.

### 2.4 Method 3: CSI recalculated via both rescaled PPV and Sens

There is also a method to recalculate CSI using not only rescaled PPV, as in Method 1, but also rescaled Sens.

Bayes formula may be used to calculate different values of NPV across the range of P values:

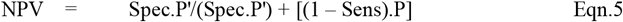

This allows recalculation of Sens at different P values using the equivalence shown by Kraemer, such that:^21^

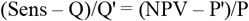

Rearranging, values for Sens at a fixed Q may be calculated at variable P:^1^

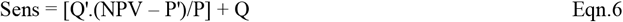

Hence this approach requires the application of Eqn.5 and Eqn.6 to the base data (Spec = 0.707; Q = 0.387 at optimal MACE cut-off of ≤20/30) to recalculate NPV and Sens respectively.

With the rescaled Sens and the previously rescaled PPV (Table 1), it is then possible to recalculate CSI (Eqn.1). Results were displayed in a table and graphically.

**Table 1:**
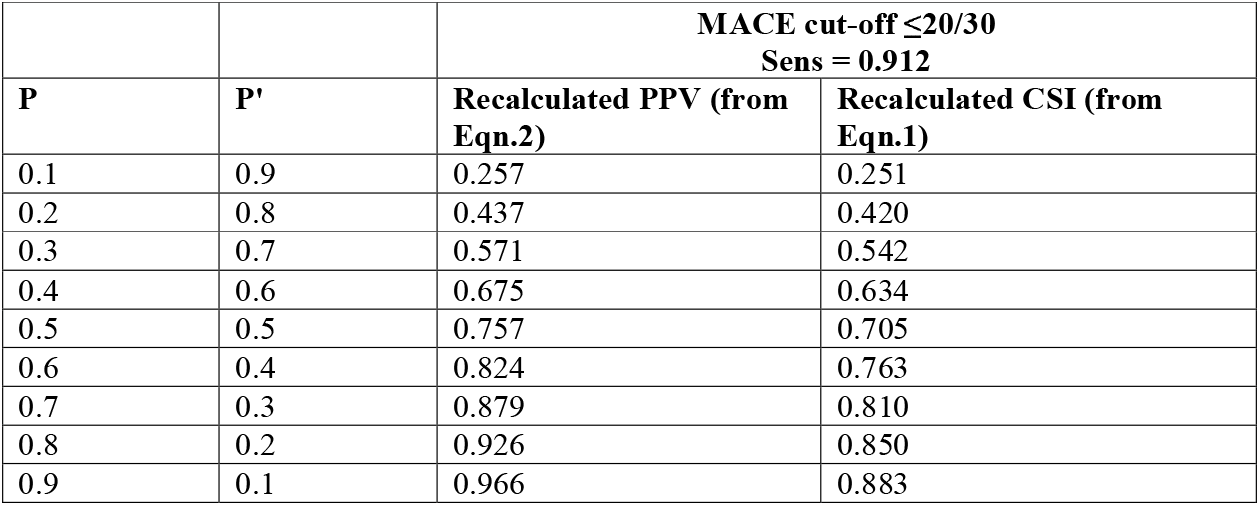
Values of PPV and CSI for dementia diagnosis at fixed value of Q (MACE cut-off of ≤20/30) at various prevalence levels.

## 3. Results

### 3.1 Method 1: CSI recalculated via Bayes formula for PPV

Using Bayes formula (Eqn.2), both the recalculated values of PPV and CSI increased with increasing P (Table 1; Figure 1A). This confirms the expectation evident in Bayes formula that CSI, like PPV, is proportional to P in this formulation. This implies that the highest values of CSI will occur when P is high.

**Figure 1:**
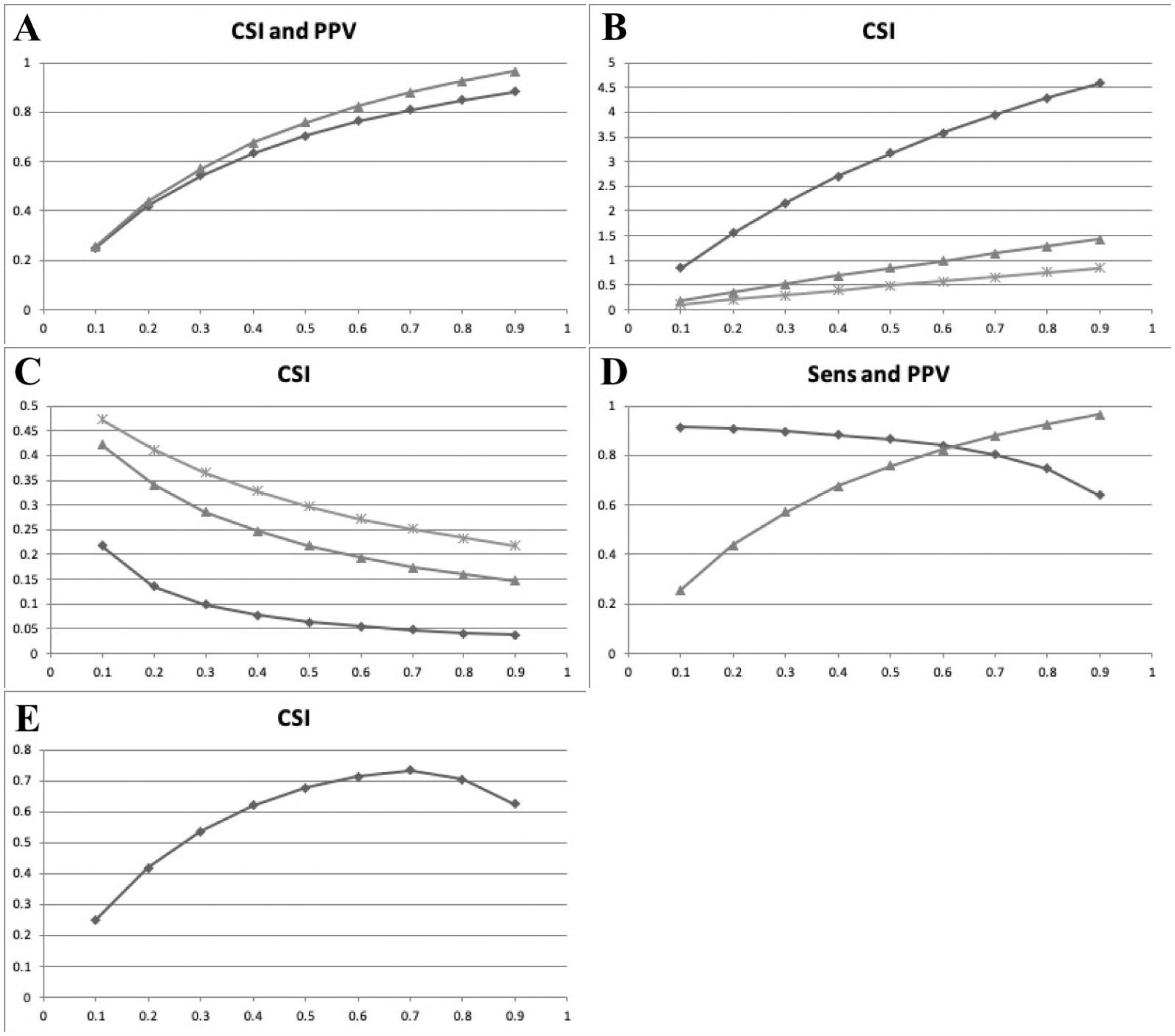
Panel of line graphs showing study results. **(A)** Plot of CSI (♦) and PPV (▲) (y axis) for dementia diagnosis at fixed Q (Q = 0.387; MACE cut-off ≤20/30) versus prevalence P (x axis) calculated by sequential application of Eqn.2 (Bayes formula) and Eqn.1 **(B)** Plot of CSI (y axis) for dementia diagnosis at fixed Sens (0.912) and variable Q = 0.1 (♦), = 0.5 (▲), = 0.9 (*) versus prevalence P (x axis) calculated using Eqn.3 **(C)** Plot of CSI (y axis) for dementia diagnosis at fixed PPV (0.356) and variable Q = 0.1 (♦), = 0.5 (▲), = 0.9 (*) versus prevalence P (x axis) calculated using Eqn.4 **(D)** Plot of Sens (♦) and PPV (▲) (y axis) for dementia diagnosis at fixed Q (Q = 0.387, MACE cut-off ≤20/30) versus prevalence P (x axis) calculated respectively by application of Eqn.6 and Eqn.2 **(E)** Plot of CSI (y axis) for dementia diagnosis at fixed Q (Q = 0.387, MACE cut-off ≤20/30) versus prevalence P (x axis), combining rescaled Sens and PPV (Figure 1D)

### 3.1 Method 2: CSI recalculated via its relation to Sens, PPV, P, and Q

Using Eqn.3 (fixed Sens value), CSI increased with increasing P (Tables 2, 3, and 4, 3^rd^ column; Figure 1B). This implies that, with a fixed Sens, the highest values of CSI will occur when P is high.

**Table 2:** Values of CSI for dementia diagnosis at fixed value of Q = 0.1 and either Sens (0.912) or PPV (0.356) at various prevalence levels.

| **P** | **P + Q** | **CSI (Eqn.3) Sens = 0.912** | **CSI (Eqn.4) PPV = 0.356** |
| --- | --- | --- | --- |
| 0.1 | 0.2 | 0.838 | 0.217 |
| 0.2 | 0.3 | 1.55 | 0.135 |
| 0.3 | 0.4 | 2.16 | 0.098 |
| 0.4 | 0.5 | 2.70 | 0.077 |
| 0.5 | 0.6 | 3.17 | 0.063 |
| 0.6 | 0.7 | 3.58 | 0.054 |
| 0.7 | 0.8 | 3.95 | 0.047 |
| 0.8 | 0.9 | 4.28 | 0.041 |
| 0.9 | 1.0 | 4.58 | 0.037 |

**Table 3:** Values of CSI for dementia diagnosis at fixed value of Q = 0.5 and either Sens (0.912) or PPV (0.356) at various prevalence levels.

| **P** | **P + Q** | **CSI (Eqn.3) Sens = 0.912** | **CSI (Eqn.4) PPV = 0.356** |
| --- | --- | --- | --- |
| 0.1 | 0.6 | 0.179 | 0.421 |
| 0.2 | 0.7 | 0.352 | 0.341 |
| 0.3 | 0.8 | 0.520 | 0.286 |
| 0.4 | 0.9 | 0.682 | 0.247 |
| 0.5 | 1.0 | 0.838 | 0.217 |
| 0.6 | 1.1 | 0.990 | 0.193 |
| 0.7 | 1.2 | 1.14 | 0.174 |
| 0.8 | 1.3 | 1.28 | 0.159 |
| 0.9 | 1.4 | 1.42 | 0.146 |

**Table 4:** Values of CSI for dementia diagnosis at fixed value of Q = 0.9 and either Sens (0.912) or PPV (0.356) at various prevalence levels.

| P | P + Q | CSI (Eqn.3) Sens = 0.912 | CSI (Eqn.4) PPV = 0.356 |
| --- | --- | --- | --- |
| 0.1 | 1.0 | 0.100 | 0.473 |
| 0.2 | 1.1 | 0.199 | 0.412 |
| 0.3 | 1.2 | 0.295 | 0.365 |
| 0.4 | 1.3 | 0.390 | 0.328 |
| 0.5 | 1.4 | 0.483 | 0.297 |
| 0.6 | 1.5 | 0.574 | 0.272 |
| 0.7 | 1.6 | 0.664 | 0.251 |
| 0.8 | 1.7 | 0.752 | 0.233 |
| 0.9 | 1.8 | 0.838 | 0.217 |

Using Eqn.4 (fixed PPV value), CSI decreased with increasing P (Tables 2, 3 and 4, 4^th^ column; Figure 1C). This implies that, with a fixed PPV, the highest value of CSI will occur when P is low.

### 3.3 Method 3: CSI recalculated via rescaled PPV and Sens

Using this method, neither PPV nor Sens is fixed, only Q. The rescaled values (Figure 1D) show Sens decreasing with increasing P (Table 5, column 4) and PPV increasing with increasing P (Table 5, column 3; and as per Table 1 and Figure 1A).

**Table 5:**
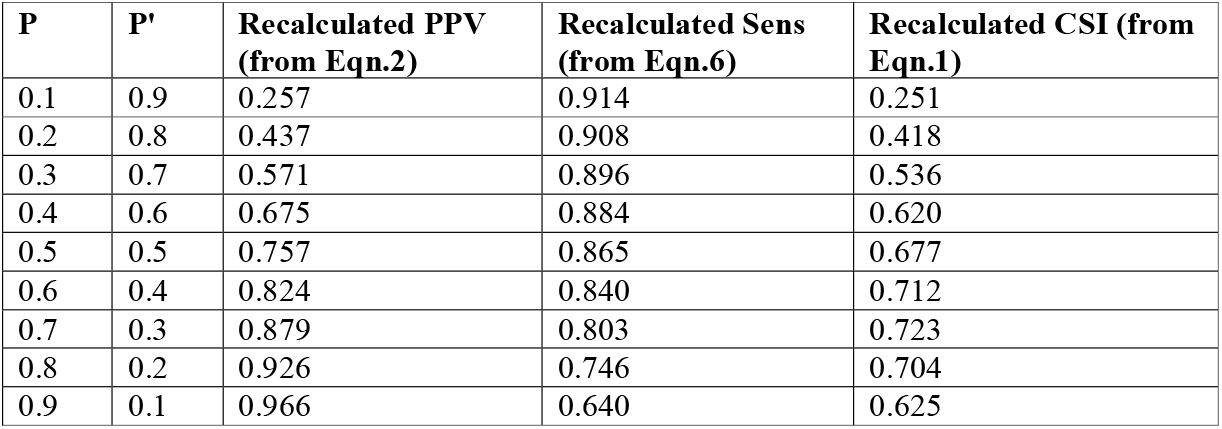
Values of recalculated PPV (as per Table 1), Sens, and CSI for dementia diagnosis at various prevalence levels.

Combining these rescaled values as per Eqn.1, CSI showed a concave curve when plotted against P (Table 5 column 5, Figure 1E). CSI values approximated PPV at low values of P (as in Figure 1A), and approximated Sens values at high values of P (compare Figures 1D and 1E).

## 4. Discussion

This study has shown that the dependence of CSI on P differs according to the method of calculation adopted.

Using either the method via Bayes formula to rescale PPV (Eqn.2) or the direct method based on Sens (Eqn.3), CSI values increased with increasing P. In these methods, the value of Sens is fixed but the product (Sens.P) varies with P. Hence CSI values increase as P increases (Figures 1A and 1B).

In contrast, using the direct method based on PPV (Eqn. 4), CSI values decrease as P increases. In this method the value of PPV is fixed, and hence the product (PPV.Q) is also fixed for each of the three chosen values of Q (Tables 2, 3, and 4, 4^th^ column). Hence the only changing variable in this method of calculation is (P + Q), which is inversely proportional to CSI (Eqn.4). This inverse relation is also expected on the basis of the observation that test Sens and PPV change in opposite directions with change in test cut-off.^6^ This change in opposite directions was empirically observed in the previous analysis of the dataset used in this study.^19^

Using the third method, in which both PPV and Sens are rescaled via Bayes formula, the relationship between CSI and P was shown to be a concave curve. This suggests that CSI will be maximal at a particular prevalence which may vary according to the particular dataset under examination. It was previously shown, using the same dataset, that another unitary measure based on Sens and PPV, the F measure (the harmonic mean of Sens and PPV) showed a concave curve when plotted against P, with a maximum value at P = 0.7 but falling away at both higher and lower values of P. The finding of maximal CSI at P = 0.7 in this dataset was previously predicted since CSI and F share a monotonic relationship.^1^ The findings suggest that, at least in this cohort, CSI values follow PPV at low values of P, and follow Sens at high values of P, but this needs further investigation in other patient cohorts.

This concave relationship is simply a reflection of the fact that CSI is dependent on both P and Q, as per Eqn.3 and Eqn.4. Just as paired outcome measures may be dependent on either P (PPV, NPV, and their complements) or Q (Sens, Spec, and their complements), so unitary measures are often functions of both P and Q. This is the case not only for CSI but also for F measure, Youden index (Y), predictive summary index (PSI), Matthews’ correlation coefficient (MCC), and the harmonic mean of Y and PSI (HMYPSI) (Table 6). All showed concave relationships to P in this dataset.^1^

**Table 6:** Summary of dependence of unitary measures on P and Q.

| Unitary measure | Dependence on P and Q |
| --- | --- |
| Critical success index (CSI) | $CSI = 1/[(P + Q)/Sens.P] - 1$<br>$CSI = 1/[(P + Q)/PPV.Q] - 1$ |
| F measure (F) | $F = 2.Sens.P/(Q + P)$<br>$F = 2.PPV.Q/(Q + P)$ |
| Youden index (Y) | $Y = (Sens - Q)/P'$<br>$Y = (Spec - Q')/P$<br>$Y = (Q - Q^2/P - P^2).PSI$ |
| Predictive summary index (PSI) | $PSI = (PPV - P)/Q'$<br>$PSI = (NPV - P')/Q$<br>$PSI = (P - P^2/Q - Q^2).Y$ |
| Matthews' correlation coefficient (MCC) | $MCC = \sqrt{(P - P^2/Q - Q^2).Y}$<br>$MCC = \sqrt{(Q - Q^2/P - P^2).PSI}$ |
| Harmonic mean of Y and PSI (HMYPSI) | $HMYPSI = 2/(1/Y).[1 + (Q - Q^2)/(P - P^2)]$<br>$HMYPSI = 2/(1/PSI).[1 + (P - P^2)/(Q - Q^2)]$ |

Hence, we suggest that there is no simple answer to the question of how CSI is dependent on P, other than that it is, and this depends on the method of calculation chosen to examine the relationship. In real-world situations, the dependence of CSI on P is not, and cannot be, independent of Q. Thus, conclusions based on outcome values of CSI (and indeed F) may be dataset-specific, and not easily translated or generalised to other situations, as is recognised to be necessarily the case for PPV. Moreover, pragmatically this is also the case for Sens since, although it is algebraically unrelated to P as a strictly columnar ratio in the 2x2 contingency table, it will vary according to the heterogeneity of clinical populations (ditto Spec),^22^ as is implied in the dependence of the Youden index on P (Table 6).

## Data Availability

Base data are available from the authors of the original study.19

## Declarations of interest

None of the authors have any conflict of interests to disclose.

## Funding

This research received no external funding.

## Data availability statement

Base data are available from the authors of the original study.^19^

## Notes

### Competing Interest Statement

The authors have declared no competing interest.

### Author Declarations

Subjects gave informed consent and the study was approved by the institute's committee on human research (Walton Centre for Neurology and Neurosurgery Approval: N 310).

